# Evaluation of HEART score, GRACE 2.0 score, and TIMI score for Risk Stratification in Acute Coronary Syndrome: A Single-Centre Pilot Study in a Sri Lankan Population

**DOI:** 10.1101/2025.11.05.25339505

**Authors:** C Welhenge, KM Fernando, JN Fernando, PC Deshapriya, MLD Lekamge, A Kasturiratne, AP De Silva

## Abstract

**Objectives:** HEART score is widely used for risk stratification, in patients with chest pain, to rule out acute coronary syndrome (ACS). Although limited, it has been validated in several countries throughout the world. TIMI and GRACE 2.0 scores are recommended for prognostication in patients with ACS. This study aimed to explore the effectiveness of HEART score, in a Sri Lankan population, for risk stratification, in patients presenting with chest pain and to compare its performance with GRACE 2.0 and TIMI scores.

**Methods:** Data was collected from 74 patients presenting to the emergency department at a tertiary care centre in Sri Lanka. HEART, GRACE 2.0 and TIMI scores were calculated for each patient separately. The predictive accuracy of the HEART score with a diagnosis of ACS and the occurrence of major adverse cardiac events (MACE) at 6 weeks was assessed and compared with TIMI and GRACE 2.0. scores.

**Results:** Area under receiver operating curve (AUC-ROC) for HEART, GRACE 2.0 and TIMI, for a diagnosis of ACS, were 0.889 (95% CI: 0.8171 - 0.9609), 0.805 (95% CI: 0.6758 - 0.9349), 0.812 (95% CI: 0.6961 - 0.9278) respectively. Delong’s test did not indicate a significant pairwise difference in scores. AUC-ROC for occurrence of MACE were 0.905 (95% CI: 0.8437 - 0.9669), 0.721 (95% CI: 0.5934 – 0.8493) and 0.767 (95% CI: 0.6467 – 0.888), for HEART, GRACE 2.0 and TIMI scores respectively, and these differences were statistically significant.

**Conclusions:** HEART score is an effective score which can be used in Sri Lankans, in predicting the probability of ACS, and MACE within 6 weeks, in patients presenting with chest pain. It is comparable to GRACE 2.0 and TIMI scores in diagnostic accuracy while it is superior to GRACE and TIMI scores in predictive accuracy for short term risk of MACE.

## INTRODUCTION

Chest pain accounts for majority of emergency medical admissions globally, including Sri Lanka (1,2). However, only about 20% of these patients are ultimately diagnosed with acute coronary syndrome (ACS), which requires immediate treatment and inward care. Delays in diagnosis of ACS, referred to as “rule out delays” contribute to overcrowding at the emergency department (ED) (3).

Risk stratification of these patients is required for early identification of patients who requires urgent and more aggressive treatment (4). Many patients presenting with chest pain are low risk and can be safely discharged and followed up as outpatients (5). Identification of patients at high risk helps the treating physicians in reducing the morbidity and mortality as well as better utilization of the limited resources in resource limited settings (4). International guidelines recommend the use of risk stratification tools in patients with chest pain due to their superiority over clinical assessment used alone and their benefits in patient outcomes (6). Expert cardiology panels recommend the utilization of dedicated risk scores for discriminating high risk patients for diagnostic and therapeutic decision making (7-9).

The HEART score was developed in 2008, for risk stratification, in patients with chest pain. It categorizes patients into low, intermediate or high risk, according to the short-term risk of major adverse cardiac events (MACE). These major cardiac events include acute myocardial infarction, the need for percutaneous coronary interventions (PCI) or coronary artery bypass grafting (CABG) and death within six weeks (5). The use of this score has been recommended as a quick, easy to use, reliable predictor of ACS at the ED as a triage tool (1). Many studies throughout the years have validated the HEART score for various populations including those in Netherland, United Kingdom, United States and South Korea (1,10 -13). A study conducted in India has validated the HEART score for Indian population, suggesting relevance for the broader South Asian region (13).

Global Registry of Acute Coronary Events (GRACE) score is a score published in 2003, to assess the in-hospital mortality of patients with ACS (13). GRACE 2.0, a more accurate version of the score was developed later, for better risk stratification and prediction of the risk of death in the acute setting as well in the long term (six months post discharge) (14). Both scores were developed and externally validated extensively in multiple regions throughout the world, including in South Asia (15-17). A GRACE score cut off, of 140 has been recommended to identify the high risk NSTE-ACS patients who require urgent coronary interventions in the European society of Cardiology (ECS), American heart association (AHA) and American college of Cardiology (ACC) guidelines.

Thrombolysis in Myocardial Infarction (TIMI) is another score, which was published in 2000. It was originally developed in patients with NSTEMI (Non-ST elevated myocardial infarction)/UA (Unstable angina) for prognostication and therapeutic decision making (18). It has also been validated in countries throughout the world, including South Asia, but less extensively when compared with the GRACE score (19-22). Although developed for use in patients with NSTEMI/UA, the score has demonstrated consistent performance in patients with potential ACS in the emergency setting (23). Nonetheless, all these scores have yet to undergo validation for the Sri Lankan population.

The purpose of this study was to assess the effectiveness of the HEART score, at predicting the probability of ACS and early risk stratification, in patients presenting with chest pain in a Sri Lankan healthcare setting. Additionally, it sought to evaluate the diagnostic and predictive performance of the HEART score in comparison with the extensively validated TIMI and GRACE scores.

## METHODS

### Participants

This study was conducted at a single tertiary care centre in Sri Lanka from July to December 2024. Data was collected from 74 adult patients presenting to the ED with chest pain. All consecutive patients above 18 years complaining of chest pain, who consented to join the study was included. Exclusion criteria comprised pregnant and post-partum patients (up to 3 months), individuals with chest pain following accidental injuries and those with known familial heart diseases. At the time of the study, PCI facilities were not available for emergency admissions at the relevant hospital. Hence, patients diagnosed with ST-elevation myocardial infarction (STEMI) who were eligible for PCI were transferred to a specialized PCI centre and were excluded from the study. However, patients who received thrombolytic therapy were included.

### Data

Data collection was performed using an interviewer-administered electronic questionnaire. Additional required data was retrieved from the bed head tickets (BHTs) and the hospital’s digital laboratory information system. Serial ECGs were taken at the ED and the ward as required for patient management. ECG interpretations made by the attending medical officers was documented and independently verified by researchers themselves and relevant sections of the questionnaire filled. hs-cTn I level, and other biochemical parameters were requested from the biochemistry laboratory as a routine practice from the ED or the ward immediately upon admission. The biochemical investigations were traced from the laboratory, once available and the relevant section of the questionnaire filled.

The data collection process occurred independently of patient management, and the calculated scores were not disclosed to or used by the treating clinical team during decision-making. The patients were reviewed prior to discharge to assess them for the final diagnosis, which was made by the treating clinical team based on the history, examination, serial ECGs, troponin values and other relevant investigations. The required information was obtained from the BHTs and confirmed with the treating clinical team. Six-week follow-up data on the occurrence of MACE were collected from the patients and personal medical records.

### Data preparation

Data was collected to a spreadsheet and recoded for statistical analysis. The HEART score was calculated from the collected independent scores using R Studio software. GRACE 2.0 and TIMI scores were calculated using validated web based freely available software.

### Outcome

The diagnosis of ACS was made by the treating clinical team in accordance with the 2023 ESC guidelines for the management of acute coronary syndromes (8) and was cross checked by the research team. Based on the clinical history, ECG findings and the hs-cTn I level, ACS was further classified as unstable angina (UA), Non-ST elevated myocardial infarction (NSTEMI) and ST elevated myocardial infarction (STEMI). Occurrence of acute myocardial infarction (AMI), the need for PCI or CABG and death within six weeks of the onset of chest pain was considered as MACE. Only NSTEMI and STEMI was considered as AMI.

### Statistical analysis

Categorical data was presented using frequencies and percentages. The effectiveness of the HEART, GRACE 2.0 and TIMI scores, were assessed with, sensitivity, specificity, positive predictive value (PPV), negative predictive value (NPV), and area under the receiver operating characteristic curve (AUC-ROC), as measures of predictive accuracy. Fisher’s exact tests were used to assess the statistical significance of the HEART score in prediction of, ACS and the risk of short-term MACE. Delong’s test for two correlated ROC curves was used to compare the AUC-ROC of the different scores. A logistic regression model, Hosmer-Lemeshow test, calibration plots and Brier scores were used to assess the discrimination, calibration and performance of a multivariable regression model. All statistical analyses were performed using R Studio (Version 2025.05.0+496). Statistical significance was defined as p < 0.05 two -sided.

### Ethical approval

Ethical approval for the study was obtained from the Ethical Review Committee of the Faculty of Medicine, University of Kelaniya, Sri Lanka (FWA00013225). Written informed consent was obtained from all participants.

## RESULTS

### Participants

Data was collected from 74 patients presenting with chest pain to the ED and followed up for 6 weeks (Figure 1).

**Figure 1.**
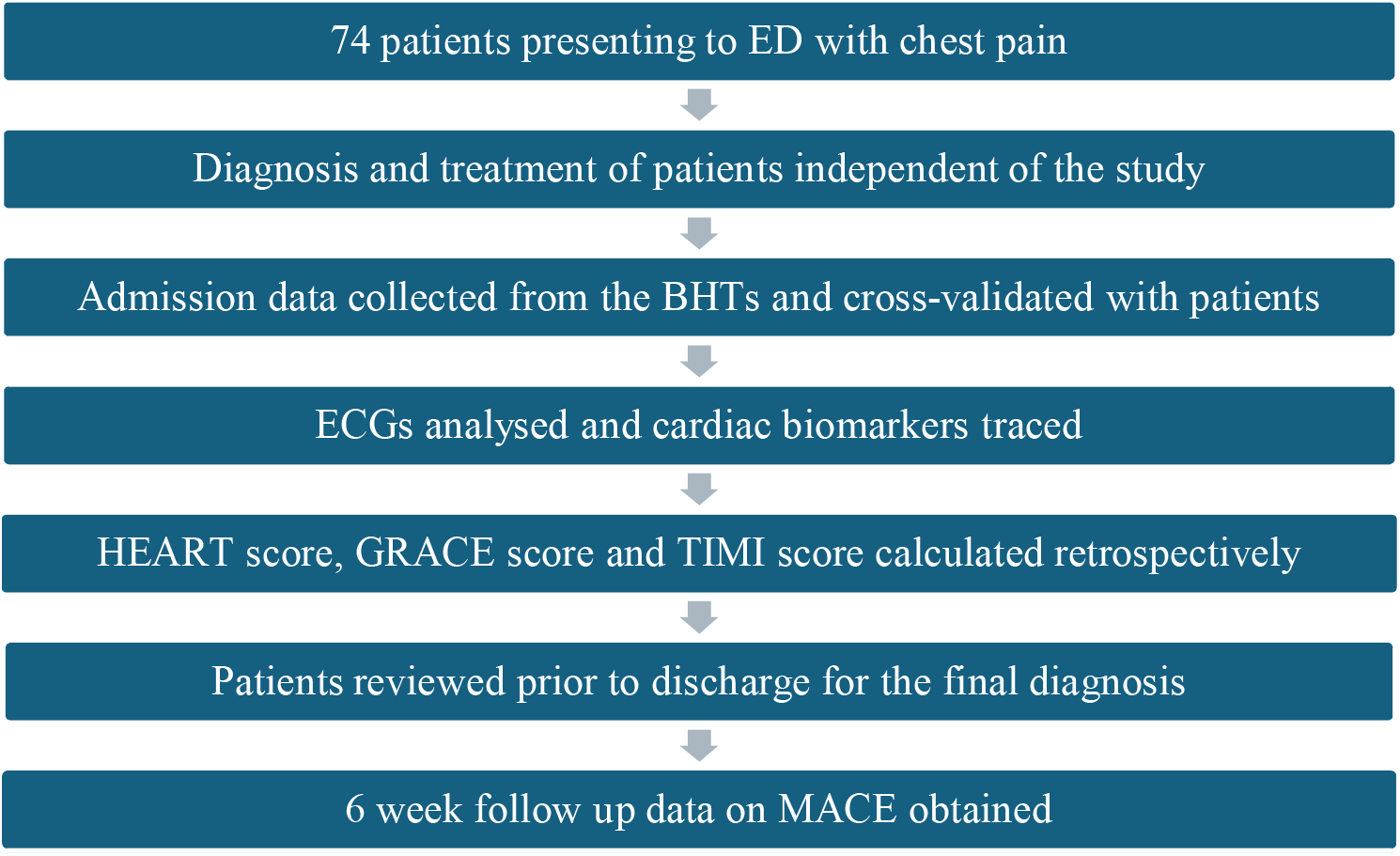

Out of the 74 patients, 42 (57%) were male and 32 (43%) were female. Patients diagnosed with unstable angina (n=9, 12.2%), NSTEMI (n=35, 47.3%) and STEMI (n=11, 14.9%) were considered to have ACS. Based on history, ECG findings and hs-cTn I value, ACS was excluded as the cause for chest pain in the rest. Among patients diagnosed with ACS at the time of discharge, MACE was observed in 48 patients (87.3%) at six weeks. In contrast, only 3 patients (15.8%) in the non-ACS group experienced MACE.

The total HEART score, GRACE 2.0 and TIMI scores were calculated retrospectively for each patient and categorized into low-risk, intermediate-risk, and high-risk groups for risk stratification (Table 1). The risk categories were compared against the diagnosis at discharge. GRACE score was not available in two patients due to missing data, and these patients were removed from analysis involving the GRACE score. The distribution of patients among risk categories between the ACS and non-ACS group is illustrated in Table 1. Fisher’s exact test to assess the statistical significance between the ACS and non-ACS group revealed p-values of 1.09 x 10^-8^, 0.002483 and 0.002676 for HEART, GRACE and TIMI scores respectively.

**Table 1.** Distribution of the HEART, GRACE and TIMI score risk categories based on the diagnosis of ACS and occurrence of MACE

| ***HEART score risk category*** | **ACS (n=55)** | **Non-ACS (n=19)** |
| --- | --- | --- |
| **Low (0-3)** | 0 | 8 |
| **Intermediate (4-6)** | 26 | 11 |
| **High (7-10)** | 29 | 0 |
| ***GRACE score risk category*** | **ACS (n=55)** | **Non-ACS (n=19)** |
| **Low ($\leq 108$)** | 16 | 14 |
| **Intermediate (109 -140)** | 27 | 5 |
| **High ($\geq 140$)** | 10 | 0 |
| ***TIMI score risk category*** | **ACS (n=55)** | **Non-ACS (n=19)** |
| **Low (0-2)** | 16 | 14 |
| **Intermediate (3-4)** | 23 | 4 |
| **High (5-7)** | 16 | 1 |
| ***HEART score risk category*** | **With MACE (n=49)** | **Without MACE (n=25)** |
| **Low (0-3)** | 0 | 8 |
| **Intermediate (4-6)** | 20 | 17 |
| **High (7-10)** | 29 | 0 |

| *GRACE 2.0 score risk category* | With MACE (n=49) | Without MACE (n=25) |
| --- | --- | --- |
| Low ($\leq 108$) | 16 | 14 |
| Intermediate (109 -140) | 23 | 9 |
| High ($\geq 140$) | 9 | 1 |
| *TIMI score risk category* | With MACE (n=49) | Without MACE (n=25) |
| Low (0-2) | 13 | 17 |
| Intermediate (3-4) | 21 | 6 |
| High (5-7) | 15 | 2 |

Abbreviations – ACS – Acute coronary syndrome, MACE - Major adverse cardiac events

All three scores were also assessed for their ability to predict MACE within six weeks (Table 1). Fisher’s exact test to assess statistical significance between risk categories of the scores and occurrence of MACE, revealed p-values of 1.965 x 10^-8^, 0.08224, 0.002206 respectively for HEART, GRACE 2.0 and TIMI scores.

In intermediate and high-risk categories of all the scores, the score was classified as positive, while in the low-risk category it was classified as negative. Cut off values (clinical cut off) and the sensitivity, specificity, PPV and NPV of each score in their ability of predicting a diagnosis of ACS and the risk of occurrence of MACE is illustrated in Table 2.

**Table 2.** Sensitivity, specificity, positive predictive value (PPV) and negative predictive value (NPV) of HEART, GRACE 2.0 and TIMI scores in predicting ACS and risk of MACE at the clinical cut off and the optimal threshold.

| Prediction of ACS |  |  |  |  |  |
| --- | --- | --- | --- | --- | --- |
| HEART |  | GRACE 2.0 |  | TIMI |  |
| *At a threshold of 4* |  | *At a Threshold of 109* |  | *At a threshold of 3* |  |
| Sensitivity | 100% | Sensitivity | 70% | Sensitivity | 71% |
| Specificity | 42% | Specificity | 74% | Specificity | 74% |
| PPV | 83% | PPV | 88% | PPV | 89% |
| NPV | 100% | NPV | 47% | NPV | 47% |
| ***Youden's optimal threshold*** | **5.5 (6)** | ***Youden's optimal threshold*** | **74** | ***Youden's optimal threshold*** | **1.5 (2)** |
| **Sensitivity** | 73% | **Sensitivity** | 96% | **Sensitivity** | 87% |
| **Specificity** | 90% | **Specificity** | 58% | **Specificity** | 58% |
| **PPV** | 95% | **PPV** | 86% | **PPV** | 86% |
| **NPV** | 53% | **NPV** | 85% | **NPV** | 61% |
| **Prediction of MACE** |  |  |  |  |  |
| **HEART** |  | **GRACE 2.0** |  | **TIMI** |  |
| ***At a threshold of 4*** |  | ***At a Threshold of 109*** |  | ***At a threshold of 3*** |  |
| **Sensitivity** | 100% | **Sensitivity** | 67% | **Sensitivity** | 74% |
| **Specificity** | 32% | **Specificity** | 58% | **Specificity** | 68% |
| **PPV** | 74% | **PPV** | 76% | **PPV** | 82% |
| **NPV** | 100% | **NPV** | 47% | **NPV** | 57% |
| ***Youden's optimal threshold*** | **5.5 (6)** | ***Youden's optimal threshold*** | **112.5 (113)** | ***Youden's optimal threshold*** | **1.5 (2)** |
| **Sensitivity** | 80% | **Sensitivity** | 65% | **Sensitivity** | 90% |
| **Specificity** | 88% | **Specificity** | 71% | **Specificity** | 52% |
| **PPV** | 93% | **PPV** | 82% | **PPV** | 79% |
| **NPV** | 69% | **NPV** | 50% | **NPV** | 72% |

*Abbreviations – ACS – Acute coronary syndrome, PPV – Positive predictive value, NPV – Negative predictive value.*

ROC curves were developed to compare the three scores for a diagnosis of ACS and predicting risk of MACE separately. The AUC-ROC of the three scores are illustrated in Table 3. The AUC-ROC of the scores were compared against each other with the Delong’s test. Compared to GRACE 2.0 score and TIMI score, HEART score had a higher AUC for the diagnosis of ACS as well as in predicting risk of MACE. The variations in the AUC-ROCs were not statistically significant, apart from the differences between the HEART score and GRACE 2.0, and the TMI score in predicting MACE. The ROC curves are illustrated in Figure 2. The Youden’s index was also calculated for each score from the ROC curves to demonstrate the optimal threshold for each score and the sensitivity, specificity, PPV and NPV of each score was calculated at this optimal threshold (Table 2).

**Table 3.**
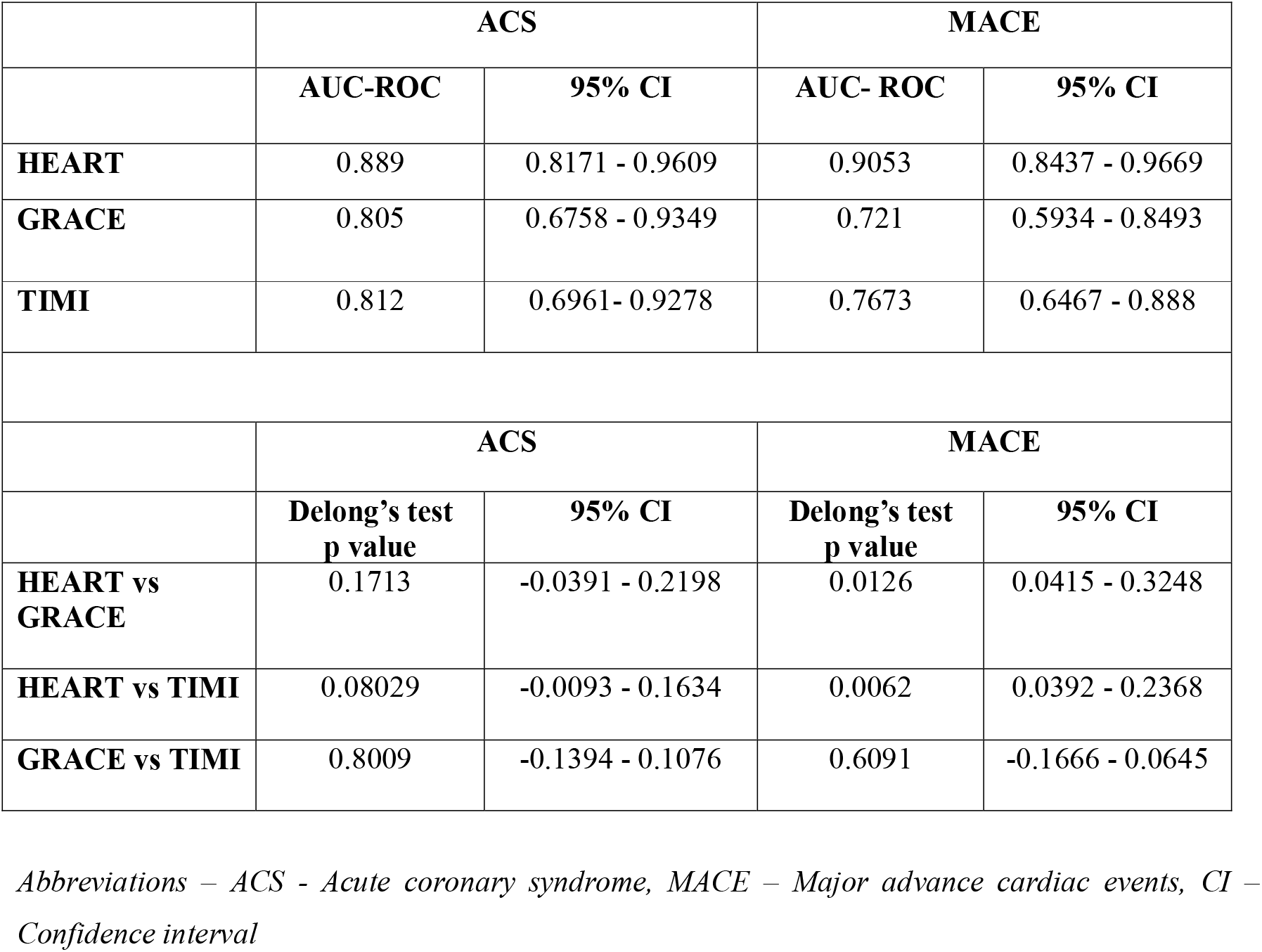
AUC-ROC of HEART score, GRACE 2.0 score and TIMI score for a diagnosis of ACS and predicting risk of MACE and statistical significance of difference between the ROC curves (DeLong’s method)

**Figure 2.**
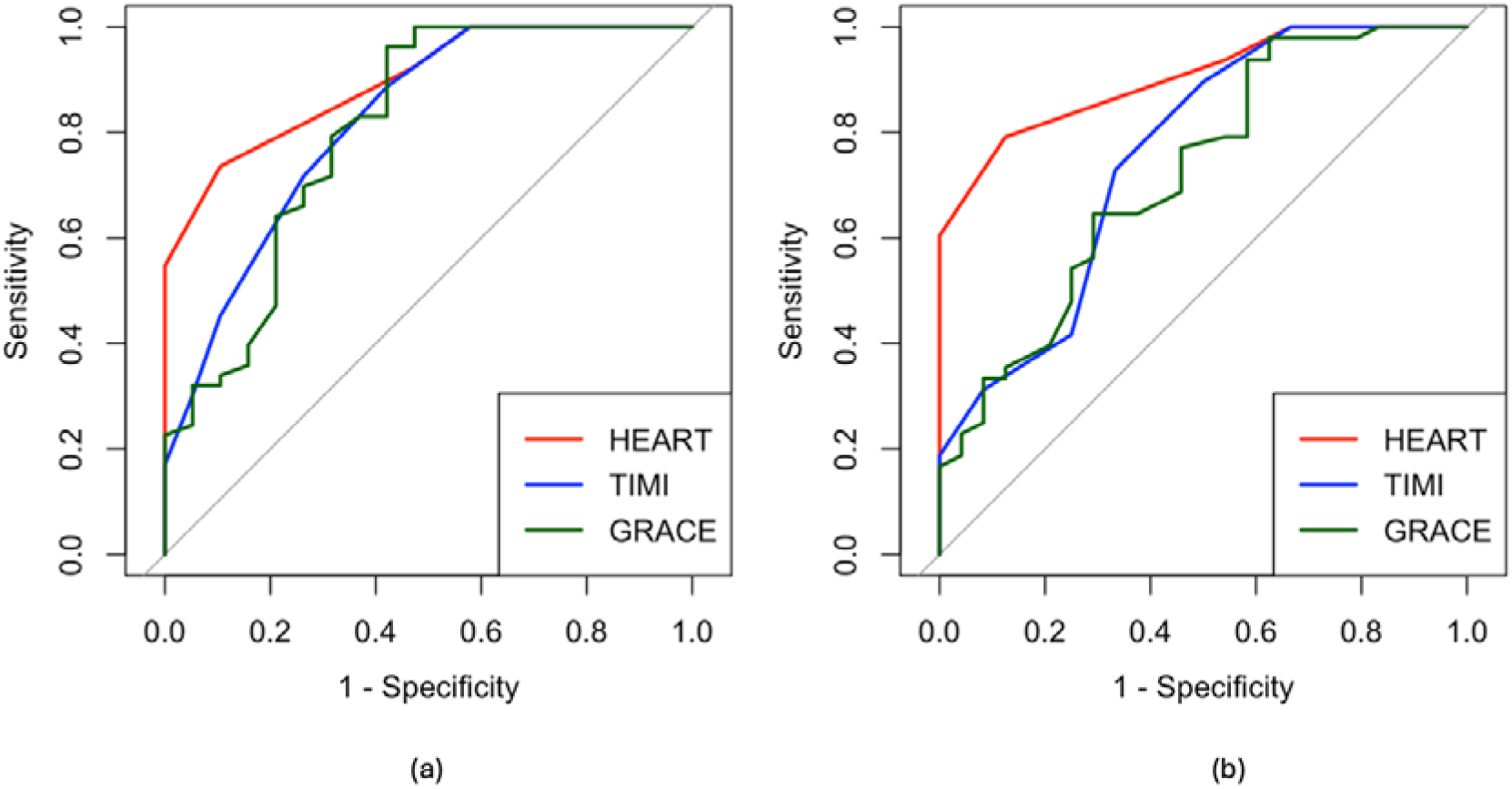
ROC curves (N=72) of HEART, GRACE 2.0 and TIMI scores (a) for a diagnosis of ACS (b) for the occurrence of MACE at 6 weeks

A multivariable logistic regression model incorporating the HEART, TIMI, and GRACE scores was used to predict the probability of ACS and occurrence of MACE (Table 4). Model performance was evaluated using AUC, calibration plots (Figure 3), and Brier scores. The AUCs of the multivariable logistic regression models for ACS and MACE prediction were 0.896 and 0.905, respectively. The Hosmer-Lemeshow goodness of fit test showed good calibration (p=0.079 and 0.399, respectively), and calibration plots using 1000 bootstrap resamples showed good alignment of predicted and observed risks. Brier scores (ACS=0.10 and MACE=0.12) were low for both models.

**Table 4.** Multivariable logistic regression model incorporating HEART, TIMI and GRACE scores to predict the probability of ACS and occurrence of MACE

| Predictor | Estimate ($\beta$) | Standard error | z-value | p-value | VIF |
| --- | --- | --- | --- | --- | --- |
| **ACS model** |  |  |  |  |  |
| Intercept | -8.115 | 2.650 | -3.06 | 0.002 | - |
| HEART score | 1.206 | 0.431 | 2.80 | 0.005 | 1.24 |
| TIMI score | -0.110 | 0.321 | -0.34 | 0.732 | 1.44 |
| GRACE score | 0.029 | 0.016 | 1.80 | 0.072 | 1.19 |
| **MACE model** |  |  |  |  |  |
| Intercept | -7.991 | 2.498 | -3.20 | 0.001 | - |
| HEART score | 1.547 | 0.446 | 3.47 | <0.001 | 1.43 |
| TIMI score | -0.311 | 0.286 | -1.09 | 0.277 | 1.64 |
| GRACE score | 0.0099 | 0.0147 | 0.67 | 0.500 | 1.20 |

Abbreviations – VIF - Variance Inflation Factor, ACS – Acute coronary syndrome, MACE – Major advance cardiac events.

**Figure 5.**
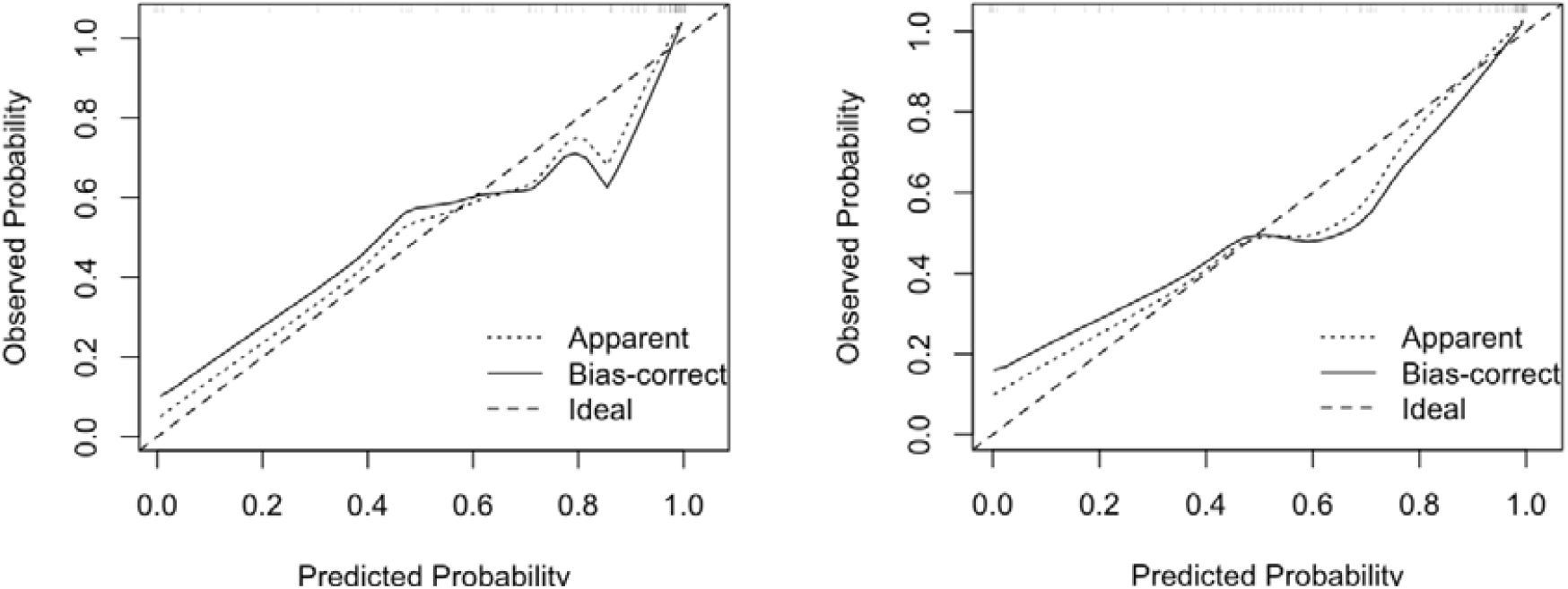
Calibration plots for predicting the probability of (a) ACS (b) MACE

## DISCUSSION

### Interpretation

Analysis of the distribution of risk categories of the three scores based on the diagnosis of ACS and occurrence of MACE demonstrated that, HEART score was the best score in precisely identifying low risk and high-risk patients. The HEART score demonstrated strong diagnostic performance, with an AUC of 0.889 for the diagnosis of ACS. The optimal threshold of the score was found to be 6. At this threshold, a sensitivity of 72.7% with a PPV of 83.3%, shows that the HEART score can be used to complement a diagnosis of ACS. A NPV of 100% with a specificity of 89.5% at the optimal threshold indicates that the score has an excellent predictive power in ruling out ACS. Further, the HEART score demonstrated excellent discriminatory power in risk prediction for six-week MACE, with an AUC of 0.9053. At the optimal threshold of 6, the score accurately identified patients at high risk of MACE (PPV = 93%) while maintaining its power in safely identifying patients at low risk of MACE (Sensitivity=80%, Specificity=88%).

Few studies over the years have attempted to evaluate the diagnostic and risk stratification accuracy of GRACE score in patients with ACS (24, 25), although it was originally intended for mortality prediction (13). Our results indicated a strong diagnostic performance of the GRACE 2.0 score for a diagnosis of ACS (AUC-ROC=0.805). At the optimal threshold of 74, the score was best for ruling in ACS (Sensitivity=96%, PPV=86%). But it can be used to safely rule out ACS as well (NPV=85%). TIMI score was similarly developed for the purpose of predicting mortality and the risk of ischemic events in patients diagnosed with ACS (18). Few studies have evaluated the applicability of the score in patients with undifferentiated chest pain (23,26). With an AUC-ROC of 0.812 for a diagnosis of ACS, our results demonstrated that TIMI score, has a good diagnostic performance in ACS, at an optimal threshold of 2 (Sensitivity=87%, PPV=86%). Given the lack of statistical significance in the difference of AUC-ROCs of the three scores, their diagnostic performance is comparable.

AUC-ROC of 0.721 and 0.7673 respectively for the GRACE 2.0 and TIMI scores, demonstrates that these scores have good discriminatory power, in risk prediction for occurrence of MACE at six weeks. At an optimal threshold of 113, which is different to the one used for predicting ACS, GRACE 2.0 showed moderate predictive accuracy (Sensitivity=65%, PPV=82%) of MACE at six weeks. TIMI score was better at identifying patients at high risk of MACE (Sensitivity=90%, PPV=79%) at an optimal threshold of 2. However, HEART score outperformed GRACE and TIMI scores, when AUCs of the three scores were compared, and the difference was statistically significant.

The multivariable logistic regression models incorporating all three scores, for ACS and MACE prediction demonstrated good discrimination (ACS=0.896, MACE=0.905). The HEART score was the only statistically significant independent predictor of ACS and MACE. Specifically, for each one-point increase in the HEART score, the odds of having ACS increased by approximately 3.4 times (OR=3.353, p=0.005), and the odds of experiencing a MACE increased by about 4.7 times (OR=4.711, p<0.001). The TIMI and GRACE scores were not independently associated with either outcome in the presence of the HEART score. No significant multicollinearity was detected (VIFs < 2 for all predictors). The Hosmer-Lemeshow test showed good calibration (ACS: p=0.079, MACE: p=0.399), suggesting good agreement between observed and predicted probabilities and calibration plots confirmed accurate agreement between predicted and observed risks. Low Brier scores (ACS=0.10, MACE=0.12) further supported the strong overall performance of the model.

### Limitations

The tertiary care centre where this study was conducted was not a designated primary PCI centre at the time of data collection. Hence the patients diagnosed with STEMI who required urgent PCI were transferred to external facilities and were not included in the study group. Although current guidelines recommend a pharmaco-invasive strategy for patients diagnosed with STEMI, in the Sri Lankan healthcare setup, where resources are limited, not all patients who receive thrombolysis proceed to coronary angiogram routinely. In addition, CABG facilities are limited, often leading to long wait times beyond the study’s follow-up period. Therefore, the actual incidence of MACE might be much higher than reported in this study.

The sample size of 74 patients, with 66% (n=49) experiencing MACE, was sufficient for pairwise AUC comparisons between the scores, and for external validation of predictive models. High event prevalence allowed stable multivariable modelling with adequate events-per-variable. Sample size calculations based on Hanley & McNeil and Obuchowski’s methods confirmed that ≥50 participants would be adequate to detect differences in AUC≥0.1 with 80% power at α=0.05. Nonetheless, a larger, multicentre study would enhance statistical power and improve the generalizability of the findings across the broader Sri Lankan Population.

### Usability of the model in the context of current care

Although the HEART score is already in use, within emergency care set up in Sri Lanka, it has not been formally validated for the local population. This study provides evidence supporting its validity among Sri Lankans. By utilizing HEART score at the EDs, ACS can be ruled out rapidly in low-risk patients presenting with chest pain and hence provide the rationale for early discharge of the low-risk patients from the ED, paving the way for reducing the overcrowding at the ED as well as the wards. Use of the GRACE 2.0 score in the ED may not be practical considering complex nature of the score as well as the requirement for serum creatinine. However, TIMI score may be used in the ED, considering its less cumbersome nature.

In resource limited settings like in Sri Lanka, the use of risk stratification tools helps in cost effective utilization of the healthcare facilities. Given the excellent discriminative power of the HEART score to predict MACE, the HEART score can also serve as an effective communication tool between emergency physicians and cardiologists when determining urgency for referral and intervention.

Refined risk stratification of patients presenting with chest pain extends beyond the emergency department. Although GRACE 2.0 score is cumbersome to be used at an emergency setting, it can be used easily in a ward setup. Identifying high risk patients assists clinical teams in therapeutic decision making and avoiding over treatment in low-risk patients. Our results show that all three scores can be utilized in the Sri Lankan population, for this purpose. These scores assist clinical teams in, treatment escalation decisions, establishing monitoring frequencies, identifying patients with residual or persistent risk despite the standard treatment who might benefit with novel treatment options etc. Therefore, the utilization of these scores needs to be encouraged in developing countries like in Sri Lanka, especially in public healthcare facilities for the optimal utilization of the limited resources.

## Data Availability

All data produced in the present study are available upon reasonable request to the authors

## OPEN SCIENCE

### Funding

No external funding was received for the study.

### Protocol

The complete study protocol can be found in the supplementary materials.

### Registration

The study has not been registered anywhere.

### Data sharing

The data that support the findings of this study are available on request from the corresponding author.

### Code sharing

The code used for the analysis is available on request.

## PATIENT & PUBLIC INVOLVEMENT

There was no patient or public involvement in planning, design, conduct, reporting or dissemination of the study and its findings

## Abbreviations

ACS: Acute coronary syndrome
ECG: Electrocardiography
ED: Emergency department
MACE: Major adverse cardiac events
PCI: Percutaneous coronary interventions
CABG: Coronary artery bypass grafting
GRACE: Global Registry of Acute Coronary Events
NSTE-ACS: non-ST elevated acute coronary syndrome
ECS: European society of Cardiology
AHA: American heart association
ACC: American college of Cardiology
TIMI: Thrombolysis in myocardial infarction
STEMI: ST-elevation myocardial infarction
NSTEMI: non-ST elevated myocardial infarction
UA: Unstable angina
BHT: Bed head ticket
hs-cTn I: High-sensitivity cardiac troponin I
AMI: Acute myocardial infarction
PPV: Positive predictive value
NPV: Negative predictive value
AUC-ROC: area under the receiver operating characteristic curve
IHD: Ischemic heart disease
VIF: Variance Inflation Factor

